# Defining Factors that Influence vaccine-induced, cross-variant neutralizing antibodies for SARS-CoV-2 in Asians

**DOI:** 10.1101/2022.03.06.22271809

**Authors:** Yue Gu, Bhuvaneshwari D/O Shunmuganathan, Xinlei Qian, Rashi Gupta, Rebecca S.W. Tan, Mary Kozma, Kiren Purushotorman, Tanusya M. Murali, Nikki Y.J. Tan, Peter R. Preiser, Julien Lescar, Haziq Nasir, Jyoti Somani, Paul A. Tambyah, SCOPE Cohort Study Group, Kenneth G.C. Smith, Laurent Renia, Lisa F.P. Ng, David C. Lye, Barnaby E. Young, Paul A. MacAry

## Abstract

The scale and duration of neutralizing antibody responses targeting SARS-CoV-2 viral variants represents a critically important serological parameter that predicts protective immunity for COVID-19. In this study, we present longitudinal data illustrating the impact of age, sex and comorbidities on the kinetics and strength of vaccine-induced neutralizing antibody responses for key variants in an Asian volunteer cohort. We demonstrate a reduction in neutralizing antibody titres across all groups six months post-vaccination and show a marked reduction in the serological binding and neutralizing response targeting Omicron compared to other viral variants. We also highlight the increase in cross-protective neutralizing antibody responses against Omicron induced by a third dose (booster) of vaccine. These data illustrate how key virological factors such as immune escape mutation combined with host factors such as age and sex of the vaccinated individuals influence the strength and duration of cross-protective serological immunity for COVID-19.

## Introduction

The emergence of SARS-CoV-2 Variants of Concern (VOC) and Variants of Interest (VOI) underlies an urgent requirement to define robust measures of protective immunity that can be utilized to triage at-risk populations and effectively target preventative countermeasures^1-5^. The Pfizer/BioNTech mRNA vaccine BNT162b2 is a leading vaccine approved by the US Food and Drugs Agency (FDA). However, there remains a significant concern about the degree of cross-protection afforded against VOC and VOI based upon multiple reports of break-through infections in partially or fully vaccinated individuals^6-8^.

There is a strong correlation between the neutralizing antibody response and protection from symptomatic SARS-CoV-2 infection^9,10^. The Delta and the Omicron variants represent the key infective strains of SARS-CoV-2 globally. Delta was first detected in India in September 2020 and remains the dominant strain in 149 countries^11^. It is being increasingly supplanted by the more transmissible Omicron variant, first identified in South Africa November 2021 and the most transmissible variant identified thus far^12-14^. Previous studies have estimated a significantly lower level of protection against the Delta variant compared to the ancestral Wuhan-Hu-1 strain upon completion of a two-dose regime of the BNT162b2 vaccine, and neutralizing responses against Beta and Gamma variants are also significantly impaired^1,4,15,16^. Moreover, several reports employing the isolated Omicron virus and/or Omicron Spike-pseudovirus have indicated a significant reduction in neutralizing antibody titres^4,17-23^.

Given the spread and immune escape potentials of SARS-CoV-2 VOCs, there remains an urgent need to evaluate cross-protective serological responses engendered by COVID-19 vaccines. Several studies have reported weaker vaccine responses in elderly populations and in males^2,3,16,24-26^. In addition, a small number of published reports have described differential responses when the seropositive population is stratified according to ethnicity, body mass index (BMI), and pre-existing comorbidities such as hypertension^24,27-29^. The impact of ethnicity on immunogenicity needs further definition, as most published studies have focussed on vaccine responses in European populations. We describe a comprehensive analysis of the cross-neutralizing and cross-binding serological responses engendered by the BNT162b2 vaccine against key viral antigens from Wuhan-Hu-1, four principal VOCs (Alpha, Beta, Gamma, Delta) and two VOIs (Epsilon and Kappa) in an Asian cohort comprised primarily of ethnic Chinese, Malay and Indian volunteers. In addition, also evaluate the serological response in both recovered patients and a subset of the vaccinees against Omicron RBD. These data have important implications for our understanding of the key factors that influence vaccine-induced immunity for SARS-CoV-2 VOC and VOI.

## Results

### Study design

Longitudinal blood samples taken pre-vaccination, three weeks post first dose, three months post first dose (peak response), and six months post first dose (long-term response) from a total of 168 participants (medium age 48, 45.8% female) were analyzed (**Fig.1a, Supplementary Table 1**). All participants were given at least two doses of the Pfizer/BioNTech BNT162b2 vaccine at 21-day intervals. In this cohort, a subset of 27 individuals had received a third dose of same vaccine (i.e. booster dose), and plasma were taken one to three months after this dose. None of the participants were diagnosed with COVID-19 previously.

**Fig. 1.**
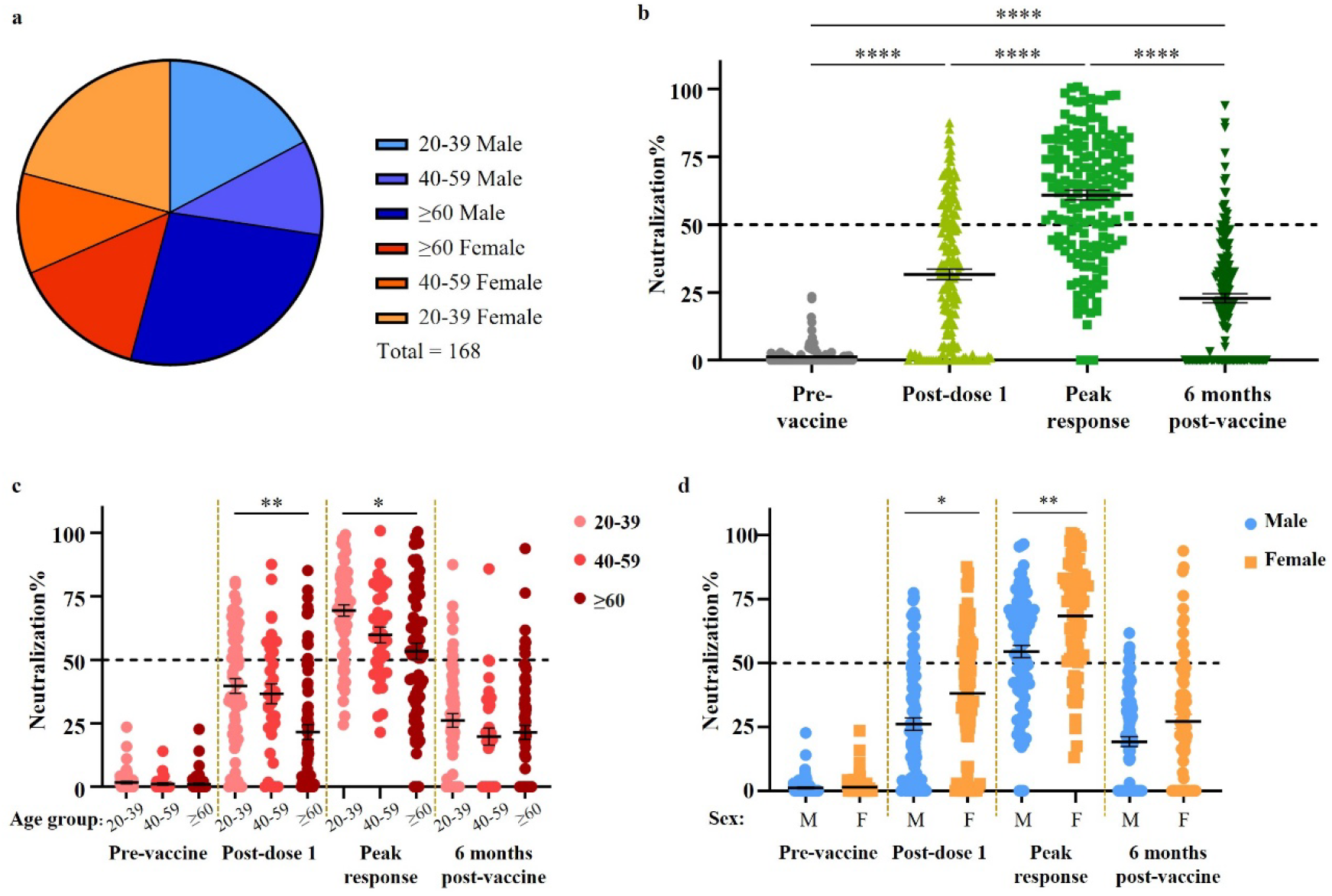
Study cohort and neutralizing response. **(a)** Demographic distribution of cohort. **(b)** Neutralizing response at four timepoints assessed by PVNT, analyzed by Kruskal-Wallis test followed by Dunn’s test. Multiple linear regression was employed to evaluate the response at each timepoint. The model contained all parameters described in this study. Results were statistically significant for **(c)** age groups and **(d)** sex. Data represented as mean ± S.E.M.,* p<0.05, ** p<0.01, **** p<0.0001.

### Serological IgG binding to SARS-CoV-2 antigens

Plasma IgG antibodies targeting SARS-CoV-2 full spike (N-terminal domain (NTD) + S1 subunit + S2 subunit), Spike-receptor binding domain (RBD) and Nucleocapsid protein (**Supplementary Fig. 1**) were measured using an enzyme-linked Immunosorbent assay (ELISA) employing fully human or humanized antibodies (for Spike, RBD and Nucleocapsid) to construct comparative standard curves and enable accurate quantitation. Anti-Nucleocapsid IgG remains negative throughout the study period for most participants (**Supplementary Fig. 2a**). Both individuals who exhibited high pre-vaccination anti-Nucleocapsid IgG were negative for anti-Spike and anti-RBD IgG (**Supplementary Fig. 2b**), indicating a high chance of cross-reactivity from previous infections by other coronaviruses. Both anti-Spike IgG and anti-RBD IgG increased significantly after two doses of the vaccine, with mean concentrations of 19.7 µg/mL and 7.4 µg/mL respectively at two months post second dose (**Supplementary Fig. 2c, d**). We observed a significant decrease in binding titers of IgG antibodies analyzed six months post-vaccination, but these titers were still higher than those measured pre-vaccination.

### Serological IgG neutralizing SARS-CoV-2 spike-pseudovirus

A SARS-CoV-2 spike pseudovirus neutralization test (PVNT) was employed to measure neutralizing activity at all time points. A significant neutralizing response was elicited by the first dose of BNT162b2 (mean 31.6% neutralization) and it improved further after the second dose with a mean of 60.9% neutralization (**Fig. 1b**). However, the neutralizing response reduced significantly to an average of 22.9% at six months post-vaccination. Vaccinated individuals below 40 years old responded strongly after a single vaccine dose compared to those aged 60 years and above. This observed difference was reduced after a second dose of vaccine. At six months post-vaccination, there was a marked decrease in the neutralizing antibody response in all age groups tested (**Fig. 1c, Supplementary Fig. 3**). In addition, we observed that females made stronger neutralizing responses after either one or two doses of vaccine (**Fig. 1d**), as previously reported.

### Development and calibration of an ELISA assay to measure neutralizing antibody responses

To augment our analyses of neutralizing antibodies based upon PVNT, we tested the ability of antibodies in volunteer plasma samples to block the interaction between angiotensin converting enzyme 2 (ACE2) and Spike-RBD. The ACE2-RBD binding inhibition enzyme-linked immunosorbent assay (ELISA) we employed for this analysis is highly predictive of the neutralizing response measured by PVNT (**Supplementary Fig. 4a-d**). Using 50% pseudovirus neutralization as the threshold to distinguish neutralizers versus non-neutralizers, the ACE2 inhibition ELISA has a sensitivity of 87.8% and specificity of 81.3% at 40% inhibition, which is employed as a threshold to define neutralizing response^30^ (**Supplementary Fig. 4e**,**f**). Moreover, we show that the observed neutralizing titres measured by this ELISA approach can be converted into predicted IC50 values through the employment of WHO reference materials and defined high, moderate, and low neutralizing samples by PVNT (**Supplementary Fig. 5**). Samples with an IC50 below 50 IU/mL exhibited less than 40% inhibition of ACE2-RBD binding in this ELISA. The tested sample set shows that over 65% ACE2 inhibition in our ELISA can predict an IC50 of above 150 IU/mL with a sensitivity of 87.5% and a specificity of 100% (**Supplementary Fig. 5b**). Using quartz crystal microbalance (QCM) technology, we observed the real-time interactions between ACE2 and RBD molecules employed in this surrogate assay, and how this interaction can be inhibited by neutralizing antibodies (**Fig. 2a, Supplementary Fig. 5d**). When pre-vaccination plasma or non-neutralizing antibody spiked plasma were allowed to interact with the immobilised RBD, subsequently injected ACE2 were able to interact with the ligand RBD, as indicated by the signal change of 15.3 Hz or 13.87 Hz respectively. In contrast, there was a smaller change in signal (11.54 Hz) when ACE2 was injected after the neutralizing antibody-spiked plasma and a minimal change (3.89 Hz) when injected after the WHO Diagnostic Calibrant (pooled convalescent plasma), implying that the ACE2 cannot interact with the coated RBD ligand in this case.

**Fig. 2.**
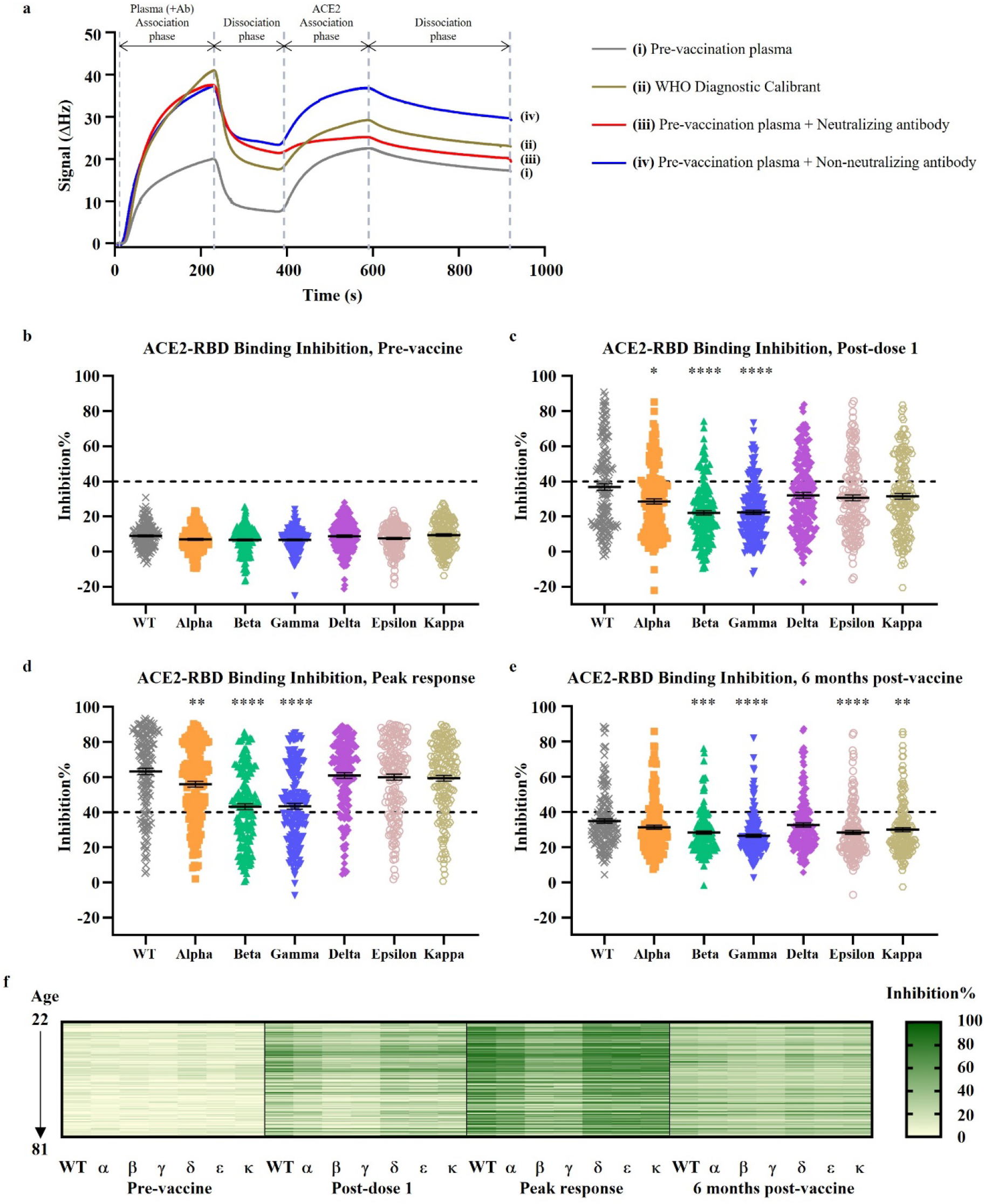
ACE2-RBD inhibitory response against VOCs and VOIs. **(a)** The effect of (i) pre-vaccination plasma, (ii) WHO diagnostic calibrant, (iii) a SARS-CoV-2 neutralizing antibody, and (iv) a SARS-CoV-2 non-neutralizing antibody on inhibiting the interaction between ACE2 and RBD were examined using quartz crystal microbalance technology. The ACE2-RBD binding inhibitory response for seven RBD variants at **(b)** pre-vaccination, **(c)** post–dose 1, **(d)** peak response post-dose 2, and **(e)** six months post-dose 1 were evaluated by ELISA. **(f)** Response at four timepoint were represented as a heatmap. Data analyzed by Kruskal-Wallis test followed by Dunn’s test. Data represented as mean ± S.E.M., * p<0.05, ** p<0.01, *** p<0.01, **** p<0.0001.

### Serological response to SARS-CoV-2 variants

In addition to the Wuhan-Hu-1 strain, neutralizing activity against the Alpha, Beta, Gamma, Delta, Epsilon and Kappa variants (**Supplementary Fig. 1**) were also evaluated using the ACE2-RBD binding inhibition. Overall, the neutralizing activity of vaccinee plasma to the Wuhan-Hu-1 strain (labelled as wild type, WT) is predictably one of the strongest amongst all variants tested. We observed significant and consistently lower levels of neutralizing activity against the Beta and Gamma strains compared to the Wuhan-Hu-1 strain at all timepoints tested until six months post-vaccination (**Fig. 2b-f**). In contrast, this trend is not observed in the binding activity to variant Spike-RBDs (**Supplementary Fig. 6**). At peak response after the second dose, the mean inhibition against all variants scored above 40%, indicating a significant degree of cross-neutralization/protection. Concordant with our observations with PVNT, the percentile inhibition of RBD-ACE2 binding for all variants also decreased at six months post-vaccination.

Females also showed a better response against all variants than males, in agreement with our PVNT data (**Supplementary Table 2, 3**). Compared to those below 40 years old, vaccine recipients aged 60 years and above exhibited a significantly lower level of antibodies that block ACE2 interactions with the Delta variant of Spike-RBD at peak response (**Supplementary Table 3**). We also observed a small but significantly stronger peak response against the Gamma variant in our Indian vaccinees compared to those of Chinese ethnicity. In addition, borderline differences against the Alpha and Beta variants were detected between these two cohorts. While needing validation in larger cohorts, this raises the possibility that significant ethnic differences in vaccine responsiveness may exist. Compared to non-smokers, a higher percentile inhibition of ACE2-RBD binding was observed in former but not active smokers (**Supplementary Table 3**). BMI and comorbidities such as hypertension and high cholesterol had no impact on the observed neutralizing titres against the VOC and VOI tested at all time points (**Supplementary Table 2-4**).

### Serological response to multiple SARS-CoV-2 variants and Omicron

Several reports on the Omicron RBD have revealed that its affinity to human ACE2 receptor is comparable to the wild type or D416G mutation RBD, despite the large number of mutations it harbours^22,31^. Here, we studied the interaction kinetics between ACE2 and RBD of all variants analyzed in this study, including Omicron (**Supplementary Fig. 7**). All equilibrium dissociation constants (K_D_) were measured in the 10^−8^ to 10^−9^ M range.

The serological response against all VOCs including Omicron was evaluated in a group of recovered convalescent individuals, and a subset of 27 vaccinees with age > 60 at three timepoints: pre-vaccination, peak response after second dose of vaccine, and one to two months after the booster dose. (**Fig. 3**) While IgG binding activity to the Omicron RBD was detected in 85.2% of vaccinated plasma samples after the second dose of vaccine (**Fig. 3b**), and in 80% of the convalescent samples (**Fig. 3d**), this was significantly lower compared to the Wuhan-Hu-1 strain and all other VOCs, except in comparison with the Beta variant in convalescent individuals. This segregation in response by variants was less prominent after the booster dose (**Fig. 3c**).

**Fig. 3.**
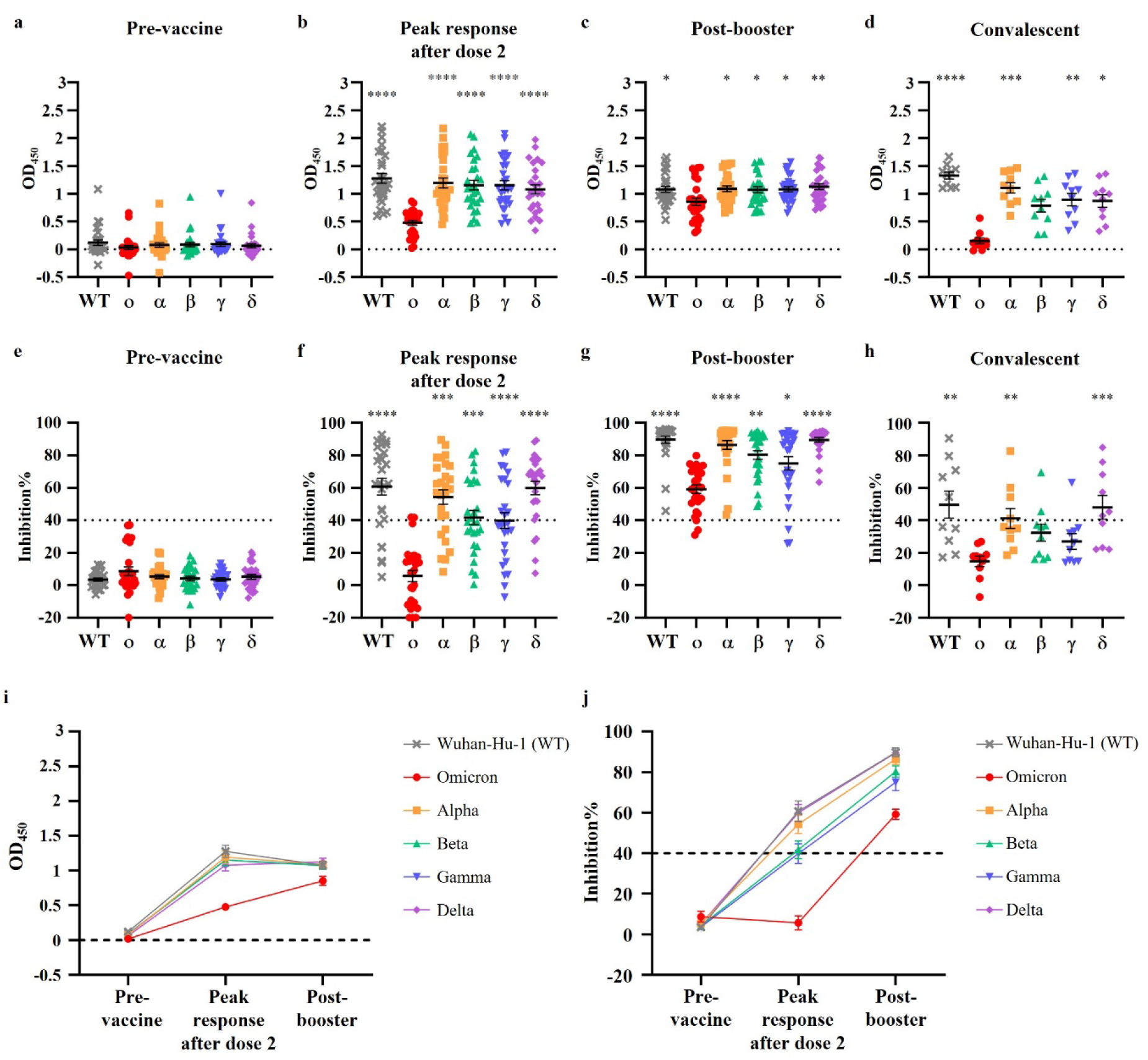
Binding activity and ACE2-RBD inhibitory response against all VOCs including Omicron in vaccinees and convalescent individuals. Serological IgG binding activity to Wuhan-Hu-1 (WT) RBD and all VOC RBDs at **(a)** pre-vaccination, **(b)** peak response after dose 2, **(c)** post-booster dose, and **(d)** post recovery from COVID-19 were measured by ELISA. **(e-h)** ACE2-RBD binding inhibitory response were also evaluated at these timepoints. **(i)** The RBD-binding activity and **(j)** ACE2-RBD inhibitory response at three timepoints were summarized for vaccinees. Response at each timepoint were analyzed by Kruskal-Wallis test followed by Dunn’s test. N=27 for (a-c, e-g, i, j). N=10 for (d, h). Data represented as mean ± S.E.M., * p<0.05, ** p<0.01, *** p<0.01, **** p<0.0001.

In concordance with the observed reduction in binding activity, neutralizing activity of post-dose 2 and convalescent plasma samples against the Omicron RBD was significantly lower than the ancestral strain and all other VOCs (**Fig. 3f, h**). Before the emergence of Omicron, immune escape effects were often reported to be the most potent in Beta and Gamma variants^32,33^. Here, we show that the post-dose 2 neutralizing response to Omicron is inferior to both Beta and Gamma variants (p < 0.001), though to a lesser extent compared to Wuhan-Hu-1, Alpha, and Delta variants (p < 0.0001). A similar trend was observed in the convalescent group (**Fig. 3h**). Again, we highlight that the neutralizing response against Omicron was much comparable to the other variants after the booster dose of vaccine (**Fig. 3g**)

Generally, we observed that the binding and neutralizing response to all the variants in the convalescent group is lower than in the vaccinated group, even without the booster dose. This is expected as many of the PCR-positive individuals included in this study only presented mild symptoms. Evidently, the booster led to a marked increase in the IgG binding response against Omicron and neutralizing response all variants (**Fig. 3i,j**).

## Discussion

The successful rollout of national vaccination programmes has been crucial in reducing mortality from pandemic COVID-19, and will continue to have an important impact once SARS-CoV-2 has become endemic. To date, 60.5% have received at least one dose of vaccine globally^34^. The long-term effectiveness of this strategy is dependent on the degree of cross-protection afforded for VOC, and the Pfizer/BioNTech BNT162b2 represents one of the key platforms employed.

In this Asian cohort, we observed a good neutralizing response for Wuhan-Hu-1 after both doses of BNT162b2, with a mean 60.9% neutralization by PVNT, translating into an average IC50 of 138.6 IU/mL. However, at peak response 30.3% (51 out of 168) of the vaccinees exhibited a neutralizing response below 50%. In addition, the mean neutralizing titres across all groups fell below 23% at six months post-vaccination, indicating a need for an additional booster dose. Our Wuhan-Hu-1 RBD-ACE2 binding inhibition ELISA result also illustrates the effectiveness of the booster dose. Among the subset of vaccinees whose post-booster plasma samples were analyzed, 22.2% (6 out of 27) showed sub-optimal ACE2 inhibition (below 40% inhibition) at peak response after the second dose. All these individuals showed an enhancement in ACE2 inhibitory response (to over 40% inhibition) after the booster dose.

In concordance with other studies, we report a significantly weaker *neutralizing response in* the group aged 60 years and above for the Delta variant, and this qualitative readout was not matched by quantitative differences in IgG binding to SARS-CoV-2 antigens. These findings are also consistent with reports of vaccine break-through infections with the Delta variant^7,8^. In addition, we observed stronger neutralizing responses among female vaccine recipients versus to males across all viral variants tested, as previously reported in Europeans^16,24,25^. We highlight that all vaccinees exhibited a significant reduction in their neutralizing response at six months post-vaccination regardless of age group (p = 0.5504), sex (p = 0.1103), or any other categories analyzed.

Long lived PCs and serum IgG correlated with protection, and the decline in antigen-specific IgG over time may be a concern. This reflects, however, only one of a number of key components contributing to effective immunological memory. Memory B cells also provide long-term memory that is qualitatively different to that reflected in serum IgG titres. Memory B cell selection results in their recognition of a relatively broad range of antigenic specificities, in contrast to long-lived PCs that are more stringently selected for affinity and specificity^35,36^. Thus, memory B cells are likely to provide a response better able to counter new VOCs such as Omicron, in a manner not reflected in serological assays. In addition to this, T cell memory also plays an important role in protection. The way in which these aspects of memory are impacted by different vaccine schedules, and by infection itself^37^, in different ethnic groups, will be important in optimising vaccine strategies.

We extended our study to compare responses to the early SARS-CoV-2 variants with those to the more recent Omicron variant. Serological responses to Omicron were markedly reduced compared to those to the ancestral Wuhan-Hu-1 strain and other VOCs, consistent with reports from other countries^17-23^. This illustrates that the immune escape effect of Omicron is greater than that of the Beta and Gamma variants. While the increased protection against Omicron from booster doses of vaccination is encouraging, follow-up studies to determine the duration of this booster effect in specific ethnic groups are required.

In summary, this study highlights the need for vigilance in monitoring the emergence of VOCs, and of assessing the efficacy of vaccination programmes in protecting against them, particularly where individuals who are above 60 years old and male are less well-protected by neutralizing antibodies. Having compared the neutralizing responses across ethnicities, we observed a small difference in a single timepoint. But overall, this difference is minor compared to age and sex, suggesting that ethnicity is not a major factor in determining the strength and durability of antibody response to the vaccine. Given the observed reduction in neutralizing antibody titres in all groups at six months post-vaccination, plus the overall augmentation in cross-protective response to all VOCs including Omicron after booster dose, this argues strongly for the implementation of vaccine booster shots that augment the degree of cross-protective responses particularly in at-risk groups.

## Methods

### Study design, ethical statement, and sample collection

The vaccinated participants were recruited under the COVID-19 PROTECT study (2012/00917). The convalescent plasma samples were collected from subjects who were diagnosed with COVID-19 by positive polymerase chain reaction (PCR) results (study 2020/00120). All participants provided written informed consent in accordance with the Declaration of Helsinki for Human Research. Ethics committee of National Healthcare Group (NHG) Domain Specific Review Board (DSRB) Singapore gave ethical approval for this work.

All vaccinated participants received two doses of the Pfizer/BioNTech BNT162b2 mRNA vaccine at 21 days apart. Four plasma samples were collected from each participant: on the day of first dose, before vaccination (i.e. pre-vaccination); on the day of second dose, before vaccination (i.e. post-dose one); three months after the first dose (i.e. peak response); and six months after the first dose. In addition, plasma sample from a fifth timepoint at one to three months after the booster dose (i.e. third dose) were collected from 27 individuals. To analyze the response to vaccination in the general population, participants with known SARS-CoV-2 infection history and participants under immunosuppressive treatments were excluded. A total of 699 plasma samples from 168 participants were included in this study.

Convalescent plasma samples (n = 10) were collected one to three months after diagnosis. All convalescent volunteers have recovered when the samples were collected.

### Expression and purification of SARS-CoV-2 antigens and receptors

SARS-CoV-2 Spike hexapro, RBD and ACE2 were purified as described elsewhere^38^. RBD variants were made using RBD as the template and KLD enzyme mix (NEB) and expressed and purified using the same method as for RBD. Primers used are listed in **Supplementary Table 5**.

Gene encoding SARS-CoV-2 Nucleocapsid (Biobasic) was cloned into pNIC28 and expressed in BL21(DE3). Cells cultured at 37 °C were induced by adding IPTG at a final concentration of 0.5 mM when the optical density (OD) reaches 0.6. After an overnight incubation at 16 °C, cells were harvested by centrifugation. Cell pellets were resuspended in 50 mM Tris, 500 mM NaCl, pH 8.0 (buffer A) and subjected to sonication. Supernatant was separated by centrifugation and incubated with cOmplete™ his tag purification resin (Roche) at 4 °C for 1 hour. After washing the resin with buffer A containing 25 mM Imidazole, Nucleocapsid was eluted with 50 mM Tris, 500 mM NaCl and 500 mM imidazole, pH8.0. Purified Nucleocapsid was concentrated using vivaspin concentrators with 30 kDa cut-off (GE healthcare) and buffer exchanged into 20 mM Tris, 300 mM NaCl, pH 8.0 using zeba™ spin desalting column with 7k MWCO (Thermo Scientific).

### Quantitative ELISA

Concentrations of anti-Spike, anti-RBD, and anti-Nucleocapsid IgG antibodies in plasma samples were estimated using quantitative ELISA^39^. Antigens were diluted to 1 µg/mL in 1x PBS and coated at 100 µL/well onto 96-well flat-bottom maxi-binding immunoplates (SPL Life Sciences #32296) by incubating at 4 °C overnight. Plate was washed three times with washing buffer (1x PBS with 0.05% Tween-20) and blocked with blocking buffer (3% bovine serum albumin in washing buffer) at 350 µL/well for 60-minutes incubation. Plasma samples were diluted 200-times and 5000-times in blocking buffer and added to the plates at 100 µL/well after plate wash step. To estimate the concentrations of anti-spike and anti-RBD antibodies, an RBD-specific human monoclonal IgG antibody named LSI-COVA-015 isolated from COVID-19 convalescent patient was diluted to a series of concentrations ranging from1 ng/mL to 1 µg/mL and added at 100 µL/well. Similarly, a nucleocapsid-specific monoclonal IgG antibody named LSI-COVANC-D generated from hybridoma cloning^40,41^ was added at the same concentrations to estimate the concentration of anti-nucleocapsid antibodies in the plasma samples. After 60-minutes incubation, plate was washed and incubated with goat anti-human IgG-HRP antibodies (Invitrogen #31413, diluted 10000-times in blocking buffer) at 100 µL/well for 50 minutes, protected from light. Plate wash step was repeated and TMB substrate (Thermo Scientific #34029) was added at 100 µL/well. After 3-minutes incubation, reaction was stopped with 100 µL/well 1 M H_2_SO_4_ and optical density at 450 nm (OD_450_) were recorded. Standard curves were constructed using the reference antibodies from 100 ng/mL to 1 ng/mL, and the concentration of antigen-specific IgG antibodies in plasma samples were calculated via interpolation.

### SARS-CoV-2 pseudotyped lentivirus production

Reverse transfection methodology was employed to generate pseudotyped viral particles expressing SARS-CoV-2 Spike proteins, using a third-generation lentivirus system. A total of 36 ×10^6^ HEK293T cells were transfected with 27 µg pMDLg/pRRE (Addgene, #12251), 13.5 µg pRSV-Rev (Addgene, #12253), 27 µg pTT5LnX-WHCoV-St19 (SARS-CoV-2 Spike) and 54 µg pHIV-Luc-ZsGreen (Addgene, #39196) using Lipofectamine 3000 transfection reagent (Invitrogen, #L3000-150) and cultured in a 37 °C, 5% CO_2_ incubator for three days. At day 4, the viral supernatant was collected and filtered through a 0.45 µm filter unit (Merck). The filtered pseudovirus supernatant was concentrated using 40% PEG 6000 via centrifugation at 1600 g for 60 minutes at 4 °C. Lenti-X p24 rapid titer kit (Takara Bio, #632200) was used to quantify the viral titers, as per manufacturer’s protocol.

### Pseudovirus neutralization test (PVNT)

ACE2 stably expressed CHO cells were seeded at a density of 5×10^4^ cells in 100 µL of complete medium [DMEM/high glucose with sodium pyruvate (Gibco, #10569010), supplemented with 10% FBS (Hyclone, #SV301160.03),10% MEM Non-essential amino acids (Gibco, #1110050), 10% geneticin (Gibco, #10131035) and 10% pencillin/streptomycin (Gibco, #15400054)], in 96-well white flat-clear bottom plates (Corning, #353377). The cells were cultured in 37 °C with humidified atmosphere at 5% CO_2_ for 24 hours. The next day, subject plasma samples were diluted to a final dilution factor of 80 with sterile 1x PBS. The diluted samples were then incubated with an equal volume of pseudovirus at a concentration of 2×10^6^ IFU/mL to achieve a total volume of 50 µL, at 37 °C for 1 hour. The pseudovirus-plasma mixture was added to the CHO-ACE2 monolayer cells and left incubated for 1 hour to allow pseudotyped viral infection. Subsequently, 150 µL of complete medium was added to each well for a further incubation of 48 hours. The cells were washed twice with sterile PBS. 100 µL of ONE-glo™ EX luciferase assay reagent (Promega, #E8130) was added to each well and the luminescence values were read on the Tecan Spark 100M. The percentage neutralization was calculated as follows:

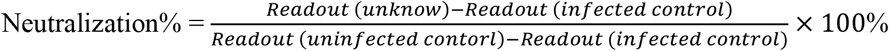

### PVNT-derived IC50 values (IU/mL) using calibrated anti-SARS-CoV-2 Immunoglobulin WHO international standard

The ID50 (Inhibitory dilution factor at 50 % neutralization) values obtained from the PVNT were converted to IC50 (inhibitory concentration at 50% neutralization) values, using the WHO international standard for anti-SARS-CoV-2 immunoglobulin (20/136). The PVNT assay was conducted as described above. The pseudovirus mixture was incubated with eight serial 5-fold dilutions of international standard (1:100 start dilution). The values were then plotted and ID50 was determined. As per WHO’s protocol, 20/136 was assigned an arbitrary value of 250 IU/ampoule (1000 IU/mL) for neutralizing activity. A calibration factor was derived based on ID50 converted to IU/mL (1000/ ID50). Following this, ID50 values were similarly obtained from nine vaccinee samples and three pooled plasma samples from the WHO reference panel (20/150, 20/148, 20/140) incubated with three serial 5-fold dilutions (1:100 start dilution). The ID50 values were then converted to IC50 (IU/mL) by multiplying the calibration factor.

### RBD variant binding ELISA

The binding ability of plasma samples to RBD variants were tested using an ELISA protocol similar to the one described earlier. RBD variants were coated at 1 µg/mL and plasma samples were tested at 100-times dilution. A negative control (100-times diluted heat-inactivated FBS) and a positive control (ACE2-Fc at 5 µg/mL in negative control) were included for each variant in each plate. Reported OD_450_ was calculated by subtracting the background OD_450_ of diluted plasma binding to blocking buffer from OD_450_ of diluted plasma binding to RBD variants.

### ACE2-RBD binding inhibition ELISA

The ability of plasma samples to inhibit the binding interaction between ACE2 and RBD variants were evaluated using a protocol similar to the RBD variant binding ELISA. Wuhan-Hu-1, Alpha, Beta, Gamma, and Delta RBD were coated at 1 µg/mL. Wuhan-Hu-1 RBD and Omicron RBD were coated at 2 µg/mL. Wuhan-Hu-1 RBD were coated at two concentrations for calibration of Omicron RBD results to account for the difference in coating concentrations. Plasma samples were tested at 5-times dilution, and the secondary antibody used was ACE2-Peroxidase (conjugated using Peroxidase-labelling kit-NH_2_, Abnova #KA0014), at 600 ng/mL for Omicron, or 300 ng/mL for all other RBD variants. A negative control (5-times diluted heat-inactivated FBS) and a positive control (ACE2-Fc at 100 µg/mL in negative control) were included for each variant in each plate. Inhibition% was calculated using the following formula:

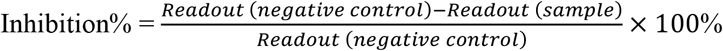

### Interaction kinetics

The binding kinetics between the human ACE2 receptor and SARS-CoV-2 RBD were measured at 22 °C using the Attana Cell 200 (Attana AB), which employs the quartz crystal microbalance technology. The Attana LNB-carboxyl sensor chips were activated by sulfo-NHS/EDC, and Wuhan-Hu-1 RBD or Omicron RBD were immobilised onto the chips to achieve a signal of about 50 Hz. Chip surface wash then quenched using ethanolamine. HBST buffer (10 mM HEPES + 150 mM NaCl + 0.005% Tween-20, adjusted to pH7.4) was used as running buffer for subsequent injections. Human ACE2-Fc molecules were added as the analyte by injecting at five different concentrations: 3.25×10^−8^ M, 6.5×10^−8^ M, 1.3×10^−7^ M, 2.6×10^−7^ M, and 5.2×10^−7^ M. Each association phase lasted for 129 seconds at a flow rate of 20 µL/minute, and dissociation was monitored for 300 seconds. Bound analyte were removed by injecting 12.5 µL of 10 mM Glycine (pH 2) over a period of 30 seconds after each analyte injection step. All concentrations were tested three times at randomized orders.

Another chip coated with Wuhan-Hu-1 RBD (signal 120 Hz after coating) was used to examine the inhibitory effect of neutralizing antibodies on RBD-ACE2 inhibition. In this experiment, 20-times diluted pre-vaccination plasma, 20-times diluted pre-vaccination plasma spiked with 1.67×10^−8^ M neutralizing antibody COVA2-39^42^, 20-times diluted pre-vaccination plasma spiked with 1.67×10^−8^ M non-neutralizing antibody LSI-COVA-15, or 20-times diluted WHO Diagnostic Calibrant (pooled convalescent plasma), were injected. After a 210s association phase and a 165s dissociation phase, 2.6×10^−7^ M ACE2 was injected without any regeneration steps. The association phase of ACE2 was 210s and the dissociation phase was observed for 330s. Regeneration of chip surface was performed using the same protocol as described above after dissociation of ACE2. All conditions were tested three times at randomized orders.

Negligible noise (<3 Hz) was detected on activated sensor chips with no ligand coated. Curve fitting using a bivalent interaction model and subsequent calculation of dissociation equilibrium constant were performed using TraceDrawer.

### Statistical analysis

Continuous demographic data were presented as median with interquartile range (IQR) while categorical demographic data and medical history information were presented as absolute number with proportion (%). Multiple linear regression was used to evaluate the effect of demographic information (age, sex, ethnicity, BMI) and medical history (smoking status, hypertension, high cholesterol) on SARS-CoV-2 antigen-specific IgG concentrations, and neutralizing antibody levels respectively. Neutralizing antibody levels and antigen-specific IgG concentrations across timepoints were evaluated using the non-parametric Kruskal-Wallis test followed by the Dunn’s test to correct for multiple comparisons. Association between the neutralization% from PVNT and inhibition% from ACE2-RBD binding inhibition ELISA was modelled using simple linear regression, and the Pearson’s correlation coefficient was also reported. Binding data and ACE2 inhibition data on RBD variants were compared to the original Wuhan-Hu-1 strains using the Kruskal-Wallis test followed by the Dunn’s test. Results on Wuhan-Hu-1 strain and other RBD variants were compared to the Omicron RBD using the Kruskal-Wallis test followed by the Dunn’s test. All statistical tests were two-tailed when applicable. All experiments were performed with three technical repeats. Mean and standard error of the mean (S.E.M.) are shown in all graphs unless otherwise stated. All analyses were performed using Graphpad Prism 9.0 and can be found in **Supplementary Data**.

### Data availability

The datasets generated during and/or analysed during the current study are available from the corresponding author on reasonable request.

## Supporting information

Supplementary Data Statistical Analyses

## Data Availability

All data produced in the present study are available upon reasonable request to the authors.

## Acknowledgement

This work was supported by the Biomedical Research Council (BMRC), A*CRUSE (Vaccine monitoring project), the A*ccelerate GAP-funded project (ACCL/19-GAP064-R20H-H) from Agency of Science, Technology and Research (A*STAR), Singapore National Medical Research Council COVID-19 Research Fund (COVID19RF-001; COVID19RF-007; COVID19RF-0008; COVID19RF-060) and A*STAR COVID-19 Research funding (H/20/04/g1/006). This study is funded by the Singapore National Medical Research Council (R-571-000-081-213, R-711-000-058-598), Ministry of Health (R-571-000-093-114), National University of Singapore (R-571-000-081-213), and the Singapore-HUJ Alliance for Research and Enterprise (R-571-002-012-592).

We thank Protein Production Platform of Nanyang Technological University for their help in making the nucleocapsid expression constructs and small-scale protein expression tests. We thank Assoc Prof. Tan Yee Joo, Department of Microbiology and Immunology, Yong Loo Lin, School of Medicine, National University of Singapore (NUS) for the ACE2 stably expressing CHO cells and plasmid encoding SARS-CoV-2 S protein for the pseudotyped lentiviral production. We thank Ms Lang Si Min for helping with the performance of experiments.

## Author contributions

Conceptualization, P.A.M.; Methodology, P.A.M., Y.G., X.Q.; Validation, Y.G., B.S.; Investigation, Y.G., B.S., X.Q., R.G., R.S.W.T., M.K., K.P., T.M.M, N.Y.J.T., H.Z., J.S., P.A.T., SCOPE Cohort Study Group; Resources, P.R.P., J.L., L.R.; Writing – Original Draft, Y.G., P.A.M.; Writing – Review and Editing, Y.G., B.S., X.Q., P.R.P., J.L., J.S., P.A.T., K.G.C.S., L.R., L.F.P.N., D.C.L., B.E.Y.; Visualization, Y.G., X.Q.; Supervision, P.A.M.; Project Administration, R.G.; Funding Acquisition, P.A.M., L.R., L.F.P.N., D.C.B., B.E.Y.

^†^SCOPE cohort study group: Siew-Wai Fong, Siti Naqiah Amrun, Yun-Shan Goh, Matthew Zi-Rui Tay, Angeline Rouers, Zi Wei Chang, Nicholas Kim-Wah Yeo, Yi-Hao Chan, Pei Xian Hor, Chiew Yee Loh, Yuling Yang, Anthony Torres Ruesta, Vanessa Neo, Wendy Yehui Chen, Estelle Yi-Wei Goh, Alice Soh-Meoy Ong, Adeline Chiew Yen Chua, Samantha Nguee, Yong Jie Tang, Weiyi Tang, Joel Xu En Wong.

## List of Supplementary Materials

**Supplementary Fig. 1. Purified SARS-CoV-2 antigens and receptor protein**.

**Supplementary Fig. 2. Quantification of serological IgG antibodies against SARS-CoV-2 antigens by ELISA**.

**Supplementary Fig. 3. Neutralizing response versus age**.

**Supplementary Fig. 4. Correlation between neutralizing response and ACE2 inhibitory response**.

**Supplementary Fig. 5. Correlation between IC50, ACE2-RBD binding inhibition response, and PVNT**.

**Supplementary Fig. 6. Binding activity of serological IgG against RBD variants**.

**Supplementary Fig. 7. Interaction kinetics between RBD and ACE2**.

**Supplementary Table 1. Demographics of study cohort**.

**Supplementary Table 2. Multiple linear regression analysis of ACE2-RBD binding inhibition response after the first dose of vaccine**.

**Supplementary Table 3. Multiple linear regression analysis of ACE2-RBD binding inhibition response at peak response**.

**Supplementary Table 4. Multiple linear regression analysis of ACE2-RBD binding inhibition response at six months post-vaccination**.

**Supplementary Table 5. Primers.**

**Supplementary Data. Statistical analyses**.

