## Supplementary Figures and Tables for "Defining Factors that Influence vaccine-induced, cross-variant neutralizing antibodies for SARS-CoV-2 in Asians"

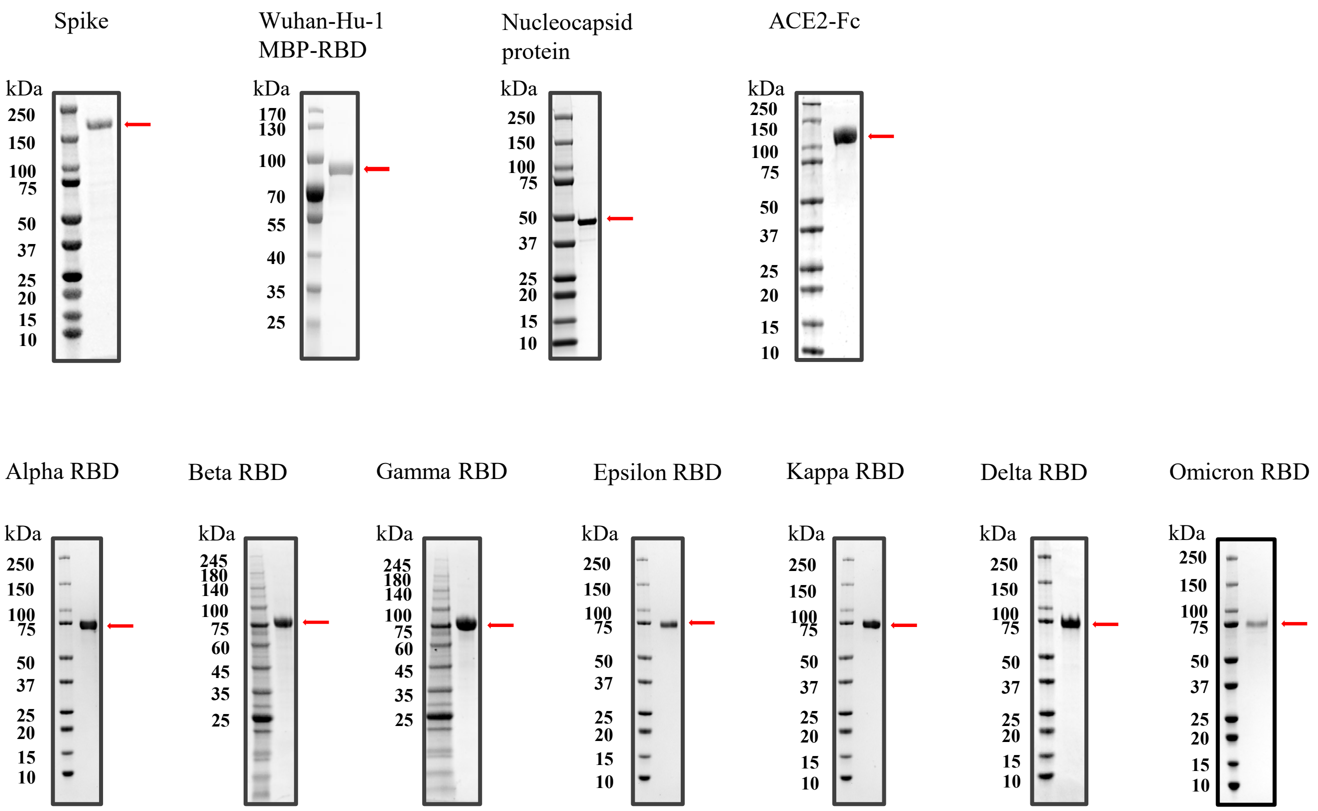


**Supplementary Fig. 1. Purified SARS-CoV-2 antigens and receptor protein.** Size of purified proteins were confirmed on SDS-PAGE gel. Proteins include Spike, RBD, and Nucleocapsid of SARS-CoV-2 Wuhan-Hu-1 strain, ACE2 receptor conjugated with IgG Fc region, and RBD of six SARS-CoV-2 variants.

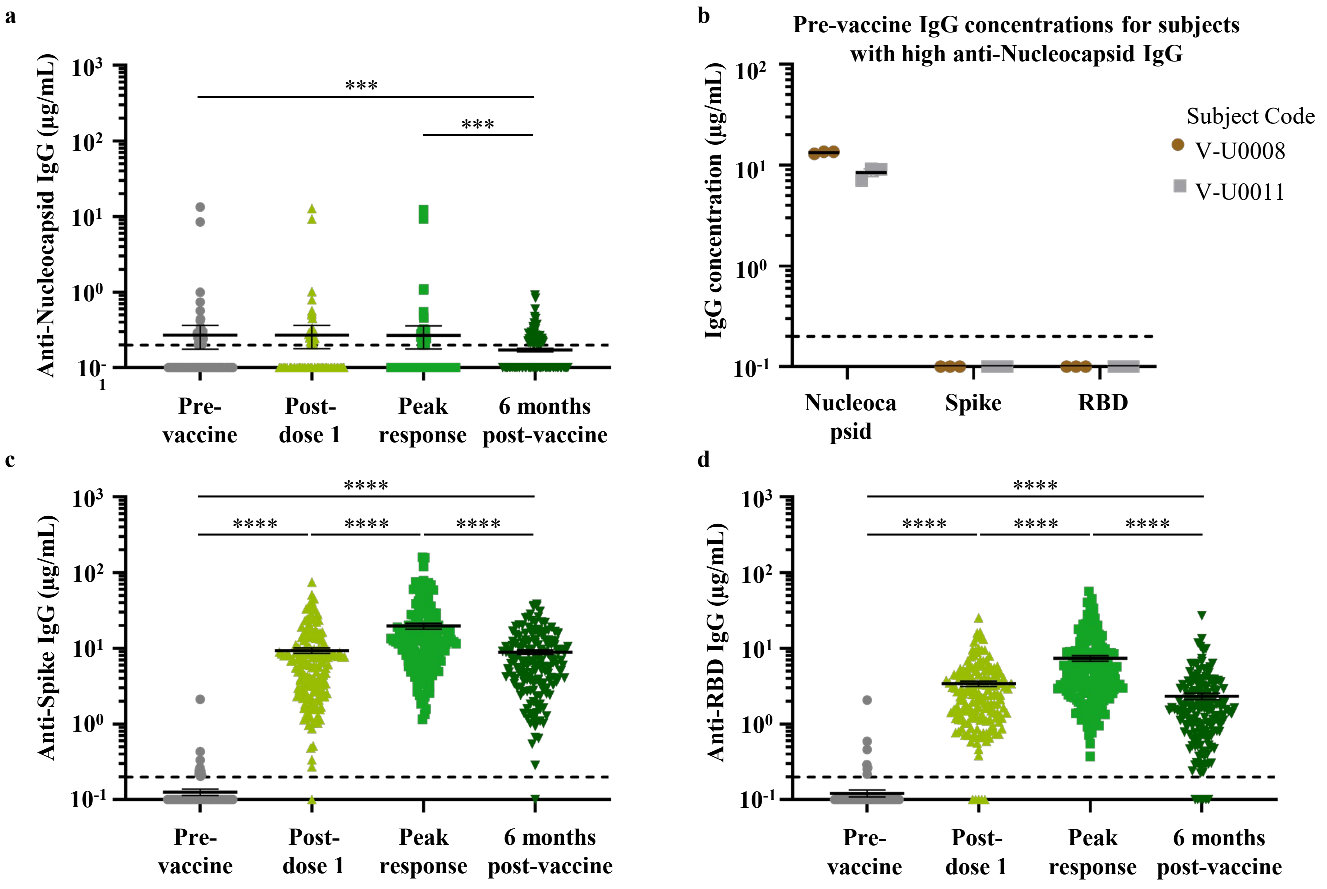


**Supplementary Fig. 2. Quantification of serological IgG antibodies against SARS-CoV-2 antigens by ELISA.** (**a**) Serological IgG antibodies against SARS-CoV-2 Nucleocapsid protein at four timepoints. (**b**) Pre-vaccination IgG levels against SARS-CoV-2 Spike and RBD were negative for two subjects with anti-Nucleocapsid IgG. Serological IgG antibodies targeting (**c**) Spike protein and (**d**) RBD were also quantified. All results were analyzed by Kruskal-Wallis test followed by Dunn's test. N=168, *** p<0.001, **** p<0.0001.

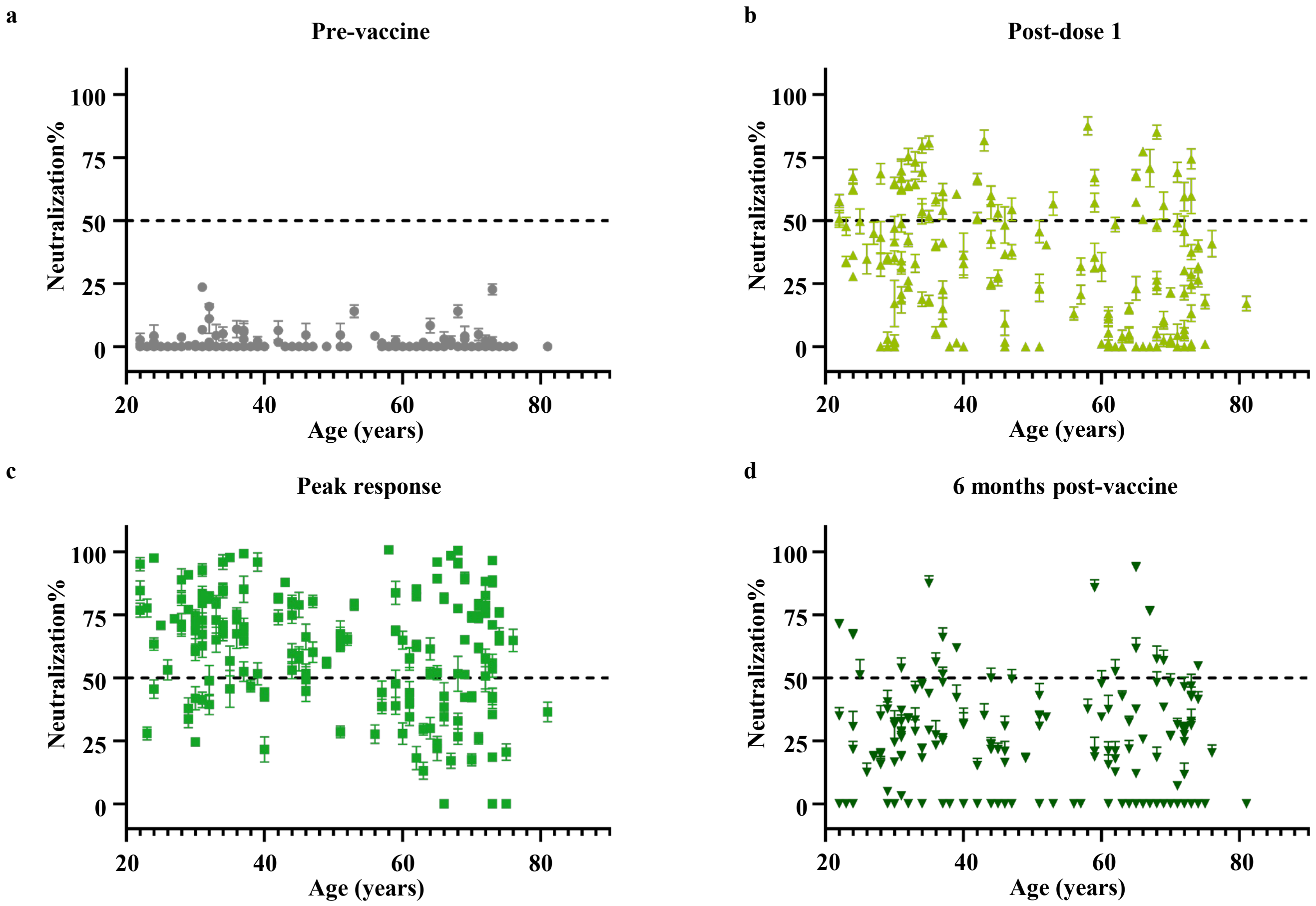


**Supplementary Fig. 3. Neutralizing response versus age.** Neutralizing response in all vaccinees were evaluated by PVNT and plotted against age (**a-d**) at four timepoints. N=168.

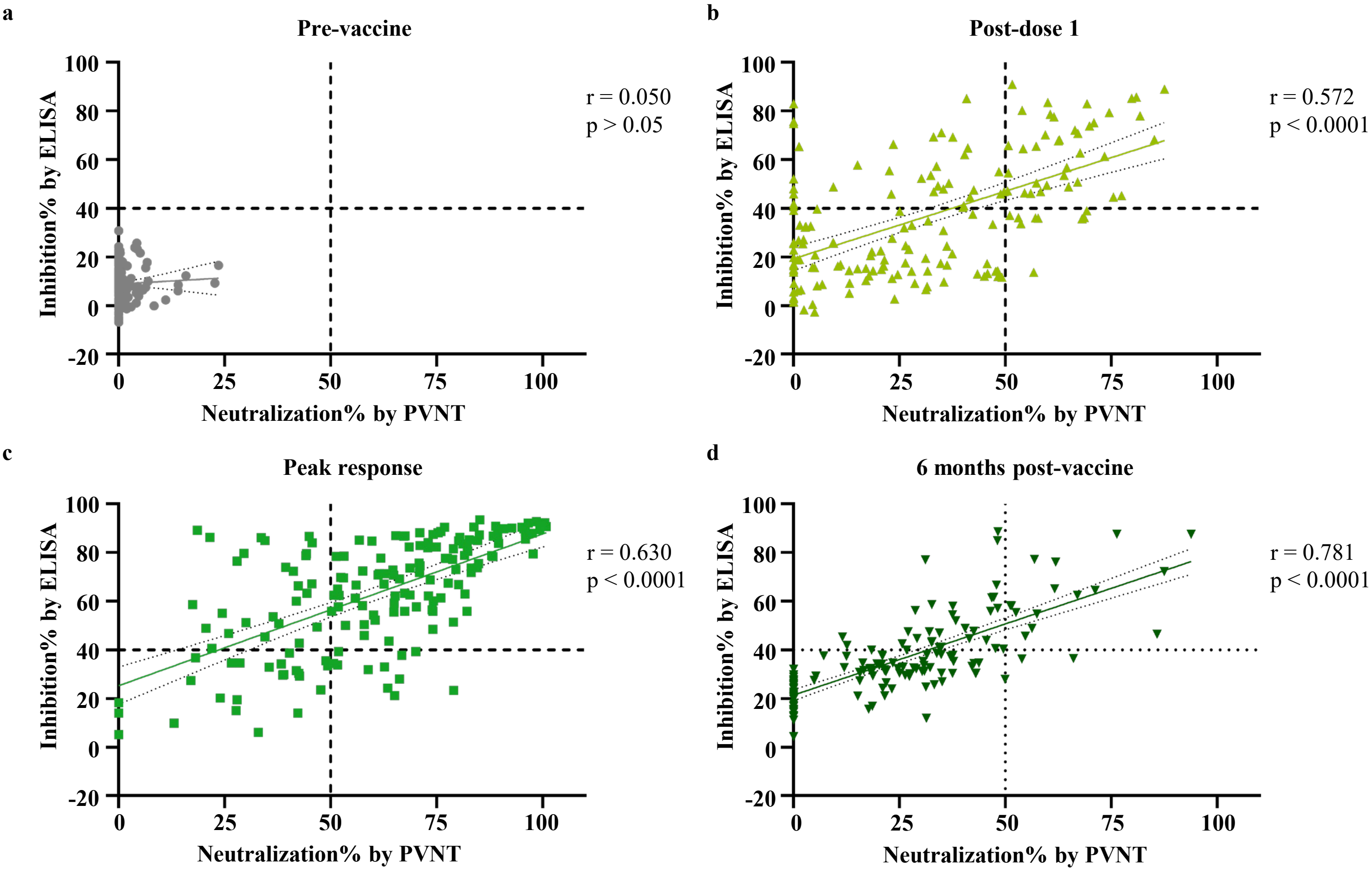


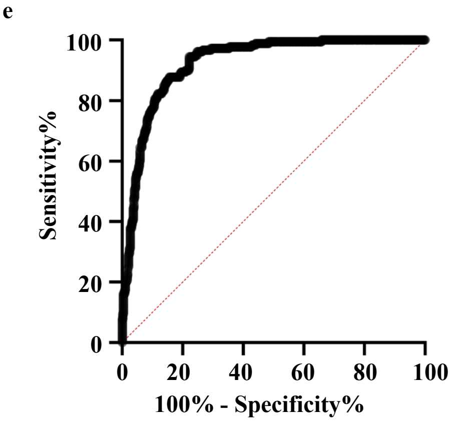


f

| | Sensitivity% | 95% CI | Specificity% | 95% CI | Likelihood ratio |
|---|---|---|---|---|---|
| > 29.80 | 97.22 | 93.66% to 98.81% | 66.87 | 62.59% to 70.88% | 2.935 |
| > 34.74 | 94.44 | 90.08% to 96.95% | 77.03 | 73.11% to 80.53% | 4.112 |
| > 39.90 | 87.78 | 82.19% to 91.79% | 81.30 | 77.62% to 84.50% | 4.694 |
| > 44.91 | 86.67 | 80.93% to 90.87% | 85.16 | 81.75% to 88.03% | 5.841 |
| > 39.90 | 80.00 | 73.56% to 85.19% | 89.23 | 86.18% to 91.67% | 7.426 |

**Supplementary Fig. 4. Correlation between neutralizing response and ACE2 inhibitory response.** Association between neutralizing response by PVNT and ACE2-RBD binding inhibition for Wuhan-Hu-1 RBD (**a-d**) at four timepoints were modelled using simple linear regression. Pearson's correlation coefficients and p-values are shown. (**e**) Using a threshold of 50% neutralization by PVNT, the predictability of ACE2-RBD binding inhibition response for defining neutralizers was evaluated with an ROC curve. (**f**) Sensitivity, specificity, and likelihood ratio close to the arbitrary threshold of 40% inhibition is shown. N=168.

a

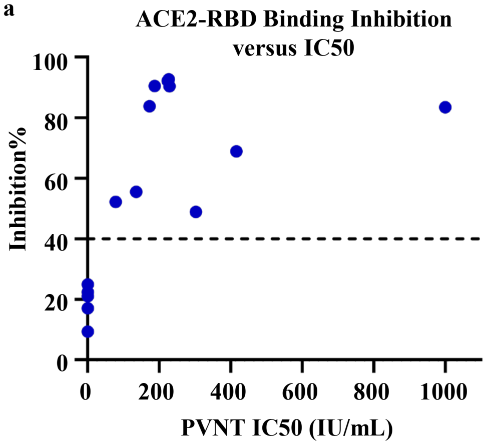


b

| | Sensitivity% | 95% CI | Specificity% | 95% CI | Likelihood ratio |
|---|---|---|---|---|---|
| > 36.92 | 100.0 | 67.56% to 100.0% | 71.43 | 35.89% to 94.92% | 3.500 |
| > 53.87 | 87.50 | 52.91% to 99.36% | 85.71 | 48.69% to 99.27% | 6.125 |
| > 62.20 | 87.50 | 52.91% to 99.36% | 100.0 | 64.57% to 100.0% | - |
| > 76.16 | 75.00 | 40.93% to 95.56% | 100.0 | 64.57% to 100.0% | - |

c

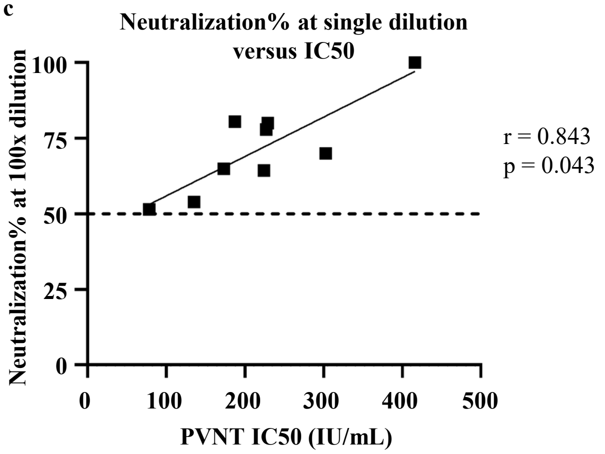


d

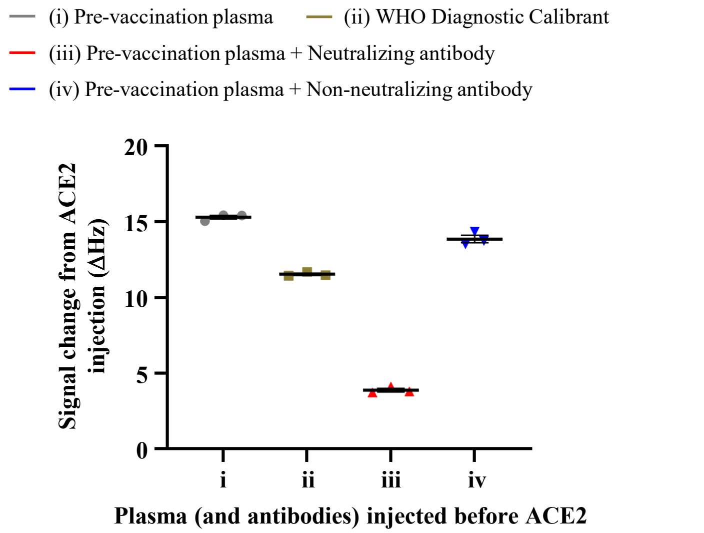


**Supplementary Fig. 5. Correlation between IC50, ACE2-RBD binding inhibition response, and PVNT.** (**a**) ACE2-RBD binding inhibition response was plotted against the PVNT-derived IC50 values after calibration with WHO international standards (n=13). IC50 values are not available (below 50 IU/mL) for all samples below the arbitrary threshold of 40% ACE2-RBD binding inhibition (n=4). (**b**) Sensitivity, specificity, and likelihood ratio of using ACE2 inhibition% as a predictor for an IC50 value of 150 IU/mL was evaluated. Figures close to the arbitrary threshold of 65% inhibition are reported. (**c**) Pearson's coefficients and p-values were calculated for the correlation between the IC50 values and neutralization% at 100x dilution of sample (n=9). (**d**) The effect of (i) pre-vaccination plasma, (ii) WHO diagnostic calibrant, (iii) a SARS-CoV-2 neutralizing antibody, and (iv) a SARS-CoV-2 non-neutralizing antibody on inhibiting the interaction between ACE2 and RBD were examined using quartz crystal microbalance technology. After RBD interaction with plasma (and antibodies), subsequent signal change during the ACE2 association phase were calculated as a parameter negatively correlated with ACE2 inhibition efficiency.

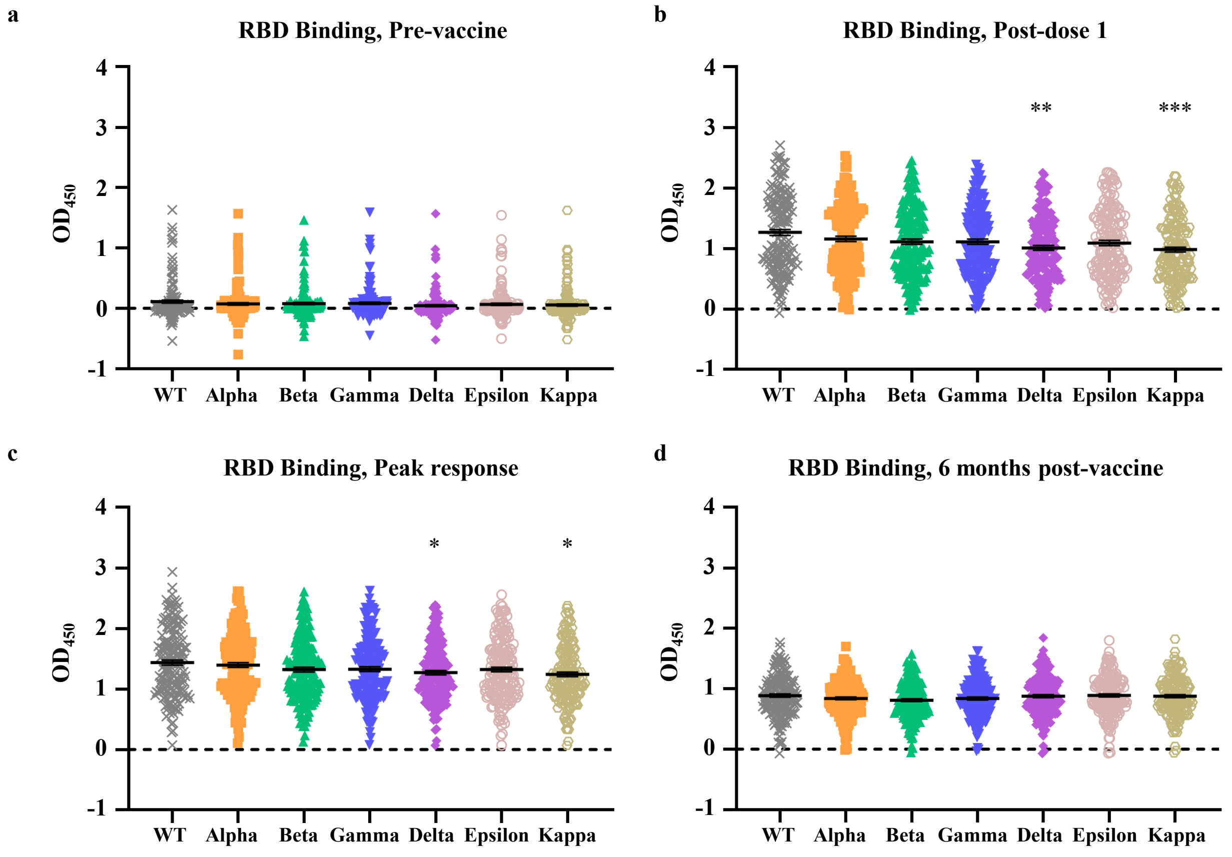


**Supplementary Fig. 6. Binding activity of serological IgG against RBD variants.** Serological IgG binding activity to seven RBD variants (**a-d**) at four timepoints were measured by ELISA. Results were analyzed by Kruskal-Wallis test followed by Dunn's test. N=168, * $p<0.05$, ** $p<0.01$, *** $p<0.001$.

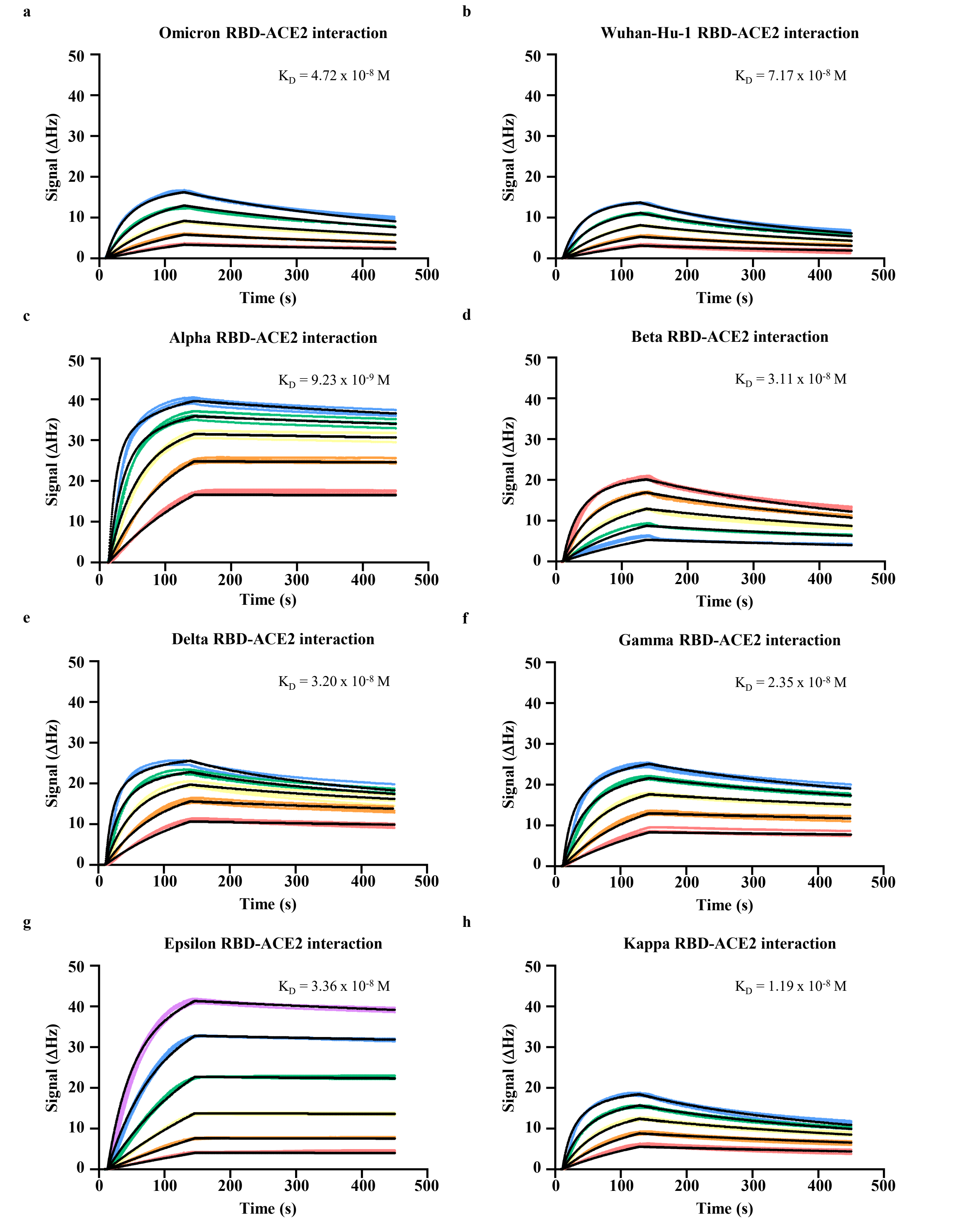


**Supplementary Fig. 7. Interaction kinetics between RBD and ACE2.** Binding kinetics between ACE2 and (**a**) Omicron RBD, (**b**) Wuhan-Hu-1 RBD, (**c**) Alpha RBD, (**d**) Beta RBD, (**e**) Delta RBD, (**f**) Gamma RBD, (**g**) Epsilon RBD, or (**h**) Kappa RBD immobilised on chips were studied using studied using quartz crystal microbalance technology. Dissociation equilibrium constants ($K_D$) was estimated based on three technical repeats.

**Supplementary Table 1. Demographics of study cohort.** Detailed information on study cohort according to age, gender, ethnicity, and comorbidities.

| | **Total (n = 168)** |
|---|---|
| Median Age (Q1-Q3) | 48 (33-67) |
| Sex (%) | 91 Male (54.2%)<br>77 Female (45.8%) |
| Ethnicity (%) | 115 Chinese (68.5%)<br>16 Malay (9.5%)<br>16 Indian (9.5%)<br>21 Others (12.5%) |
| Smoking History (%) | 23 Former smoker (13.7%)<br>16 Active smoker (9.5%)<br>129 Non-smoker (76.8%) |
| Hypertension (%) | 44 Yes (26.2%)<br>124 No (73.8%) |
| High Cholesterol (%) | 30 Yes (17.9%)<br>138 No (82.1%) |
| Mean BMI (Q1-Q3) | 24.7 (22.2-27.7) |

**Supplementary Table 2. Multiple linear regression analysis of ACE2-RBD binding inhibition response after the first dose of vaccine.** Model contained all parameters listed below and ANOVA p-values are reported. When p<0.05, parameter estimate with 95% CI and p-values are reported.

| | Wuhan-Hu-1 | Alpha | Beta | Gamma | Delta | Epsilon | Kappa |
|---|---|---|---|---|---|---|---|
| **Age** | P=0.0348 | P=0.0551 | P=0.0721 | P=0.2217 | P=0.1627 | P=0.2478 | P=0.0615 |
| >60 | -11.97 (-21.14 to -2.789), p=0.0109 | | | | | | |
| 40-59 | -3.095 (-12.34 to 6.147), p=0.5093 | | | | | | |
| **Sex** | P=0.0002 | P=0.0012 | P<0.0001 | P<0.0001 | P=0.0004 | P=0.0006 | P=0.0020 |
| Female | 14.24 (6.982 to 21.50), p=0.0002 | 9.775 (3.934 to 15.62), p=0.0012 | 10.79 (5.807 to 15.77), p<0.0001 | 10.85 (5.971 to 15.72), p<0.0001 | 12.01 (3.471 to 18.54), p=0.0004 | 11.74 (5.125 to 18.35), p=0.0006 | 10.58 (3.926 to 17.24), p=0.0020 |
| **Ethnicity** | P=0.1374 | P=0.0771 | P=0.0770 | P=0.1492 | P=0.2382 | P=0.2972 | P=0.2035 |
| Malay | | | | | | | |
| Indian | | | | | | | |
| Others | | | | | | | |
| **Smoking History** | P=0.2463 | P=0.2119 | P=0.1712 | P=0.3984 | P=0.5352 | P=0.5021 | P=0.8025 |
| Former smoker | | | | | | | |
| Active smoker | | | | | | | |
| **Hypertension** | P=0.5729 | P=0.3375 | P=0.2978 | P=0.2185 | P=0.2068 | P=0.4028 | P=0.4468 |
| Yes | | | | | | | |
| **High Cholesterol** | P=0.9466 | P=0.9859 | P=0.7319 | P=0.6066 | P=0.6252 | P=0.5724 | P=0.4369 |
| Yes | | | | | | | |
| **BMI** | P=0.3510 | P=0.1382 | P=0.1578 | P=0.1385 | P=0.1497 | P=0.1567 | P=0.0790 |

**Supplementary Table 3. Multiple linear regression analysis of ACE2-RBD binding inhibition response at peak response.** Model contained all parameters listed below and ANOVA p-values are reported. When p<0.05, parameter estimate with 95% CI and p-values are reported.

| | Wuhan-Hu-1 | Alpha | Beta | Gamma | Delta | Epsilon | Kappa |
|---|---|---|---|---|---|---|---|
| **Age** | P=0.1115 | P=0.0559 | P=0.2186 | P=0.1621 | P=0.0239 | P=0.0558 | P=0.0403 |
| >60 | | | | | -11.23 (-19.27 to -3.187), p=0.0065 | | -10.66 (-18.89 to -2.432), p=0.0114 |
| 40-59 | | | | | -5.535 (-13.63 to 2.562), p=0.1789 | | -4.186 (-12.47 to 4.103), p=0.3201 |
| **Sex** | P=0.0216 | P=0.0072 | P=0.0001 | P=0.0004 | P=0.0018 | P=0.0083 | P=0.0186 |
| Female | 8.379 (1.246 to 15.51), p=0.0216 | 9.182 (2.521 to 15.84), p=0.0072 | 13.23 (6.660 to 19.80), p=0.0001 | 12.02 (5.487 to 18.55), p=0.0004 | 10.22 (3.857 to 16.58), p=0.0018 | 9.312 (2.428 to 16.20), p=0.0083 | 7.843 (1.331 to 14.36), p=0.0186 |
| **Ethnicity** | P=0.1222 | P=0.0872 | P=0.0933 | P=0.0460 | P=0.2928 | P=0.1904 | P=0.1934 |
| Malay | | | | 5.448 (-5.537 to 16.43), p=0.3288 | | | |
| Indian | | | | 14.51 (4.176 to 24.85), p=0.0062 | | | |
| Others | | | | 4.229 (-5.590 to 14.05), p=0.3962 | | | |
| **Smoking History** | P=0.0371 | P=0.0319 | P=0.0206 | P=0.0256 | P=0.0323 | P=0.0268 | P=0.0327 |
| Former smoker | 11.17 (1.191 to 21.15), p=0.0285 | 10.80 (1.484 to 20.12), p=0.0234 | 12.20 (3.010 to 21.39), p=0.0096 | 10.65 (1.513 to 19.79), p=0.0226 | 10.21 (1.312 to 19.11), p=0.0248 | 10.70 (1.065 to 20.33), p=0.0297 | 10.65 (1.542 to 19.76), p=0.0222 |
| Active smoker | -5.732 (-17.49 to 6.032), p=0.3373 | -5.198 (-16.18 to 5.786), p=0.3514 | -3.202 (-14.04 to 7.632), p=0.5602 | -5.858 (-16.63 to 4.917), p=0.2845 | -5.139 (-15.63 to 5.352), p=0.3347 | -6.993 (-18.35 to 4.361), p=0.2256 | -4.780 (-15.52 to 5.960), p=0.3807 |
| **Hypertension** | P=0.2022 | P=0.2374 | P=0.3082 | P=0.1459 | P=0.1047 | P=0.1041 | P=0.1656 |
| Yes | | | | | | | |
| **High Cholesterol** | P=0.8012 | P=0.5201 | P=0.8895 | P=0.3450 | P=0.1786 | P=0.1415 | P=0.1188 |
| Yes | | | | | | | |
| **BMI** | P=0.7024 | P=0.8605 | P=0.8050 | P=0.7771 | P=0.7931 | P=0.8516 | P=0.9270 |

**Supplementary Table 4. Multiple linear regression analysis of ACE2-RBD binding inhibition response at six months post-vaccination.** Model contained all parameters listed below and ANOVA p-values are reported. When p<0.05, parameter estimate with 95% CI and p-values are reported.

| | Wuhan-Hu-1 | Alpha | Beta | Gamma | Delta | Epsilon | Kappa |
|---|---|---|---|---|---|---|---|
| **Age** | P=0.0970 | P=0.1713 | P=0.2339 | P=0.3003 | P=0.1352 | P=0.0708 | P=0.1627 |
| >60 | | | | | | | |
| 40-59 | | | | | | | |
| **Sex** | P=0.7434 | P=0.2694 | P=0.5628 | P=0.2395 | P=0.6458 | P=0.4480 | P=0.6576 |
| Female | | | | | | | |
| **Ethnicity** | P=0.5290 | P=0.6031 | P=0.4882 | P=0.5541 | P=0.6599 | P=0.6439 | P=0.5071 |
| Malay | | | | | | | |
| Indian | | | | | | | |
| Others | | | | | | | |
| **Smoking History** | P=0.2159 | P=0.1106 | P=0.2113 | P=0.3226 | P=0.2153 | P=0.3289 | P=0.2182 |
| Former smoker | | | | | | | |
| Active smoker | | | | | | | |
| **Hypertension** | P=0.1362 | P=0.3031 | P=0.4387 | P=0.4473 | P=0.1460 | P=0.0959 | P=0.1828 |
| Yes | | | | | | | |
| **High Cholesterol** | P=0.2618 | P=0.2341 | P=0.2988 | P=0.2166 | P=0.3299 | P=0.1774 | P=0.1484 |
| Yes | | | | | | | |
| **BMI** | P=0.3447 | P=0.3679 | P=0.1630 | P=0.1616 | P=0.0677 | P=0.1413 | P=0.1331 |

**Supplementary Table 5. Primers.** List of primers used to generate constructs for RBD variants.

| Primer | Sequence |
|---|---|
| K417N F | atattgctgattataattataaattaccagatga |
| K417 R | ttccagtttgccctggagcga |
| E484K F | aaaggttttaattgttactttccttta |
| E484K R | aacaccattacaaggtgtgct |
| A570D F | atgacactactgatgctgtccgt |
| A570D R | caatgtctctgccaaattgttgga |
| N501Y F1 | ggtgttggttaccaaccatacaga |
| N501Y R1 | gtaagtgggttggaaaccatatgattg |
| L452R F | aggtatagattgtttaggaagtctaatctc |
| L452R R | gtaattataattaccaccaaccttagaa |
| K417TF | ctattgctgattataattataaattaccagatga |
| E484K 417 R | taaaggaaagtaacaattaaaacctttaacaccattaca |
| N501Y 417 F | caatcatatggtttccaacccacttacggtgttggttac |
| E484Q F | caaggttttaattgttactttccttta |
| T478K R | aacaccattacaaggtttgct |
| E484 F | gaaggttttaattgttactttccttta |
| L452Q F1 | ggtggtaattataattaccagtatagattg |
| L452Q R1 | aaccttagaatcaagattgttagaattc |
